# Characterising associations between mental distress, mobility, and COVID-19 restrictions: a U.S. study

**DOI:** 10.64898/2026.02.26.26347164

**Authors:** Stefania Fiandrino, Saumitra Kulkarni, Paolo Cornale, Sara Ghivarello, Piero Birello, Simone Maria Parazzoli, Federico Moss, Alessandro De Gaetano, Daniele Liberatore, Jacopo D’Ignazi, Kyriaki Kalimeri, Michele Tizzani, Mattia Mazzoli

**Affiliations:** Department of Computer, Control, and Management Engineering Antonio Ruberti, Sapienza University of Rome, Rome, Italy; ISI Foundation, Turin, Italy; Network Science Institute, Northeastern University, Boston, USA; Grupo Interdisciplinar de Sistemas Complejos (GISC), Departamento de Matemáticas, Universidad Carlos III de Madrid, Leganés, Spain; Department of Network and Data Science, Central European University, Vienna, Austria; UPF, Barcelona, Spain; DTU, Copenhagen, Denmark

## Abstract

Large-scale epidemics are consistently associated with increased psychological distress and substantial changes in human mobility, yet the relationship between mental health responses and effective population mobility remains overlooked. During the COVID-19 pandemic, non-pharmaceutical interventions (NPIs) such as lockdowns and travel restrictions altered daily movement patterns while simultaneously affecting psychological well-being. Importantly, formal policy stringency alone does not fully capture realized mobility behavior, which also reflects spontaneous adaptation and adherence fatigue over time. In this study, we examine the association between self-reported mental distress and mobility recovery across the United States during the first wave of the COVID-19 pandemic. We combine state-level human mobility data derived from anonymized mobile phone records with large-scale survey data on self-reported anxiety and depression. Our analysis focuses on the U.S. states and territories from April 1 to September 1, 2020. Using fixed-effects regression models, we assess how variations in mental distress relate to deviations from pre-pandemic mobility levels, while controlling for reported COVID-19 mortality and the stringency of NPIs. We find a negative and statistically significant association between mental distress and mobility recovery: higher levels of self-reported anxiety and depression are associated with lower recovery of pre-pandemic mobility. These results indicate that psychological distress is associated with population mobility beyond what is explained by formal restrictions alone. Our findings highlight the relevance of mental health as a factor linked to behavioral responses during public health crises. Incorporating psychological well-being into the evaluation of mobility dynamics may inform more balanced public health strategies in future emergencies.

**Author summary:** During the COVID-19 pandemic, governments introduced restrictions on movement, such as stay-at-home orders and travel limits, to slow the spread of the virus. At the same time, many people experienced increased anxiety and depression. In this study, we ask whether changes in mental well-being were linked to how quickly people returned to their usual patterns of movement. Here, we focus on the first wave of the pandemic in the United States and combine mobility data and large-scale digital survey data to study the association between self-reported mental health indicators and effective mobility at the population level. By comparing states over time, we explore whether changes in mental distress were associated with changes in mobility, beyond what can be explained by public restrictions or reported deaths alone. We find that states with higher levels of reported anxiety and depression tended to show slower recovery toward normal mobility levels. This suggests that psychological well-being played an important role in shaping individual and collective responses to the pandemic, with implications for the design of future public health interventions.

## Introduction

During large-scale health crises, the association of multiple epidemic outcomes, such as fear of infection [1], financial strain [2], negative information exposure [3, 4], mobility restrictions [5, 6], and social isolation [7, 8] with psychological distress is increasingly recognized [9]. Over time, longitudinal studies indicate that such stress responses are linked to adverse health outcomes, highlighting their relevance as early indicators of preventable mental health problems. [10, 11]. The psychological burden has also been observed among caregivers and healthcare workers [12] and individuals affected by severe illness, hospitalized survivors of the severe acute respiratory syndrome (SARS) in the 2003 outbreak that reported significantly higher stress levels than individuals who had not been infected [13]. Across recent epidemics, including the mentioned 2003 SARS, the 2009 swine flu (H1N1), the 2014–2016 West African Ebola virus, and the 2015–2016 Zika virus, and the more recent 2020 coronavirus SARS-CoV-2, numerous studies have consistently reported a heightened prevalence of symptoms such as anxiety, depression, post-traumatic stress, and suicidality [14, 15]. The COVID-19 pandemic significantly affected many aspects of life [16], including social, economic, and psychological well-being. In the early stages, the absence of a vaccine prompted governments to adopt non-pharmaceutical interventions (NPIs) to contain transmission and protect public health. These measures included closure of facilities, mobility restrictions such as travel bans and stay-at-home orders. These resulted in prolonged social isolation and shifts in human mobility patterns [16–18], with non-negligible implications for psychological well-being [19] and loss of adherence to restrictions, i.e. fatigue, over time [20–22]. Given the scale and reach of this health crisis, COVID-19 offers a critical context for examining the role of mental distress responses to restrictions in shaping population travel behavior.

Studies consistently highlighted how mobility restrictions during lockdowns were linked to increased psychological distress, changes in medication use, and shifts in online behavior reflecting economic and mental health concerns [23–27]. More specific studies have analysed associations of reported mental health outcomes with stringency of restrictions, residential stationarity and reported COVID-19 cases and deaths [5, 28]. However, stringency indexes are not sufficient at describing effective population mobility as shown by the observed lack of adherence to prolonged restrictions [20, 21]. The relation between effective mobility, resulting from both imposed and spontaneous behavioral change, and mental health outcomes has been so far overlooked. This element may further clarify the complex interplay between mobility changes observed over time and population response to public health interventions.

In this study, we investigate the relationship of self-reported anxiety and depression with mobility recovery across the United States, accounting for the uncoordinated implementation and relaxation of non-pharmaceutical interventions [29], and reported COVID-19 deaths via a fixed-effects regression. This approach controls for U.S. state-specificities such as socio-economic heterogeneities and political leaning [30]. Our analysis covers 51 U.S. states and territories over the period from April 1 to September 1, 2020. We integrate human mobility data derived from mobile phone records collected by Safegraph [31] since 2019 with self-reported mental health data, specifically, indicators of anxiety, collected through the large-scale COVID-19 Trends and Impact Survey [32] administered via social media platforms.

Our findings indicate a negative and statistically significant association between self-reported mental distress and mobility recovery: higher levels of self-reported anxiety and depression are associated with lower recovery of pre-pandemic mobility levels, accounting for stringency and mortality as co-factors. This study sheds light on the association of mental distress with behavioral response to public health interventions during a health crisis. Understanding this relationship provides valuable insight into how psychological well-being can relate to mobility dynamics in the aftermath of restrictive interventions. These findings may support the design of more balanced public health strategies that account for psychological outcomes in future emergencies.

## Materials and methods

In the following section, we describe the data sources and methodology used to measure relative changes in mobility compared to pre-pandemic levels, self-reported anxiety, and contextual factors such as policy stringency and COVID-19 mortality. The analysis covers daily data from 51 U.S. states and territories, including the District of Columbia, during the initial lockdown phase of the COVID-19 pandemic in 2020. We conduct a state-level analysis, as non-pharmaceutical interventions (NPIs) were implemented by individual states rather than at the federal level. Moreover, state-level aggregation provides a more consistent and reliable basis for comparison, given potential inconsistencies in data coverage at finer spatial resolutions.

### Mobility data

We obtain the human mobility dataset from SafeGraph [31], which captures origin-destination (OD) flows across the United States based on anonymized mobile phone location data. The dataset includes daily and weekly estimates of population movements at the census tract, county, and state levels, beginning on January 1st, 2019. In this study, we focus on the daily county-level data and construct a metric representing the relative change in mobility compared to baseline pre-pandemic levels.

We examine daily mobility flows between all U.S. counties over the period from April 1 to September 1, 2020. Let *OD*_*ij*_(*t*) denote the number of trips between counties *i* and *j* on day *t*, forming a symmetric daily OD matrix. The active mobility for county *i* is defined as the total number of trips originating from that county, as reported in Eq. 1.

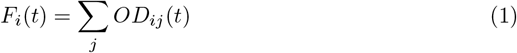

The mobility metric is aggregated to the state level by averaging *F*_*i*_(*t*) across all counties within each of the 50 U.S. states and the District of Columbia. Finally, we apply a 7-day point-centered rolling average on the time series to reduce noise from daily fluctuations. We then quantify daily mobility changes by computing the relative difference between 2020 and 2019 values for the same calendar day, using 2019 as a baseline to adjust for seasonality in mobility behavior [33] (see Eq. 2).

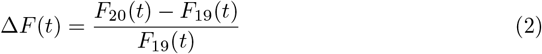

The mobility metric Δ*F* (*t*) captures the dynamic, state-level relative change in mobility and serves as a key indicator of mobility recovery trends following the abrupt decline observed during initial restriction periods. It enables us to track how, and how quickly, mobility levels rebounded over time relative to pre-pandemic norms.

### Self-reported anxiety data

To assess population-level self-reported mental distress, we used data from the COVID-19 Trends and Impact Survey, collected by the Delphi Research Group at Carnegie Mellon University in collaboration with Facebook [32]. The survey, conducted daily from April 2020 to June 2022, collected responses from a large, geographically distributed sample of U.S. residents, including self-reported measures of anxiety and depression. In this study, we focus on responses to the question regarding anxiety, specifically, whether individuals reported feeling anxious at any point during the five days preceding the survey. Answers were provided on a four-point Likert scale: “None of the time” (1), “Some of the time” (2), “Most of the time” (3), and “All of the time” (4). We derive the fraction of anxiety and depression episodes among participants by summing responses in categories 2, 3, and 4, capturing any level of reported anxiety and depression. Individual responses have been weighted by state, age, and sex to reflect population-level estimates. We computed fractions of reported mental distress at the state level. The temporal window aligns with mobility data, covering April 1 to September 1, 2020.

### COVID-19 restriction policies and mortality rates

To capture the spatial heterogeneity in policy responses during the pandemic, we include data on non-pharmaceutical interventions using the Stringency Index developed by the Oxford COVID-19 Government Response Tracker (OxCGRT) project [34]. This composite index quantifies the daily severity of government restrictions, based on nine policy indicators: school and workplace closures, cancellation of public events, restrictions on gatherings and internal movement, public transport closures, stay-at-home orders, public information campaigns, and international travel controls. The index ranges from 0 to 100, with higher values reflecting stricter policy enforcement. For epidemiological context, we incorporate daily data on confirmed COVID-19 deaths at the state level from The New York Times repository [35], which aggregates reports from state and local health agencies. We compute daily new deaths by differencing consecutive cumulative entries, and then obtain mortality rates by dividing by the state population.

### Fixed-effect panel regression model

We assess the association between mobility and self-reported mental health conditions using fixed effects panel regressions across U.S. states and territories. Specifically, we implement two separate models: one with self-reported anxiety and one with self-reported depression as the main independent variable. Both models control for COVID-19 mortality rates and policy stringency over time. The general specification of the fixed effects panel regression model is shown in Equation 3.

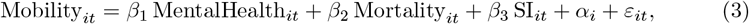

where Mobility_*it*_ denotes the mobility outflow in state *i* at time *t*; MentalHealth_*it*_ corresponds either to self-reported anxiety or depression; Mortality_*it*_ is the (log-transformed) mortality rate; and SI_*it*_ is the COVID-19 policy stringency index. The term *α*_*i*_ represents state fixed effects that control for all time-invariant characteristics of each state, and *ε*_*it*_ is the idiosyncratic error term.

Entity (state) fixed effects were included to account for unobserved, time-invariant factors that might influence baseline mobility, such as geography, demographic composition, or infrastructure. This allows the model to focus on within-state variation over time rather than cross-state differences. Because time-invariant factors are absorbed by the fixed effects, we did not include additional static covariates. We cluster standard errors at the state level to produce robust inference in the presence of serial correlation and heteroskedasticity within states. This means that observations within the same state are allowed to be correlated over time, but observations across different states are treated as independent. Therefore, clustering adjusts the standard errors to prevent underestimated uncertainty and inflated statistical significance [36]. Before estimating the association between mobility and self-reported mental health outcomes, we assessed potential multicollinearity among the explanatory variables (anxiety, depression, COVID-19 mortality, and the stringency index) using Variance Inflation Factors (VIFs). Based on this assessment, we excluded Georgia, Maine, and Oregon, which exhibited VIF values greater than 10 for at least one predictor. Across all three states, the only variable with a VIF exceeding 10 was mortality, with values ranging from 10.32 to 11.63. Consequently, the final regression models were estimated on a balanced panel of 48 states observed over 17 weeks (April to September 2020).

## Results

We first explore the temporal dynamics of the relative change in mobility during 2020 compared with 2019, averaged across all U.S. states and territories, and compare it with changes in the COVID-19 Stringency Index (Figure 1). Mobility declines sharply starting in week 11 of 2020, coinciding with the U.S. National Emergency declaration on March 13, 2020. This decline occurs in parallel with a rapid increase in the Stringency Index, indicating the widespread implementation of containment measures. During the summer, the Stringency Index gradually declines as mobility begins to recover. However, mobility remains below its 2019 baseline for the rest of the year, indicating a sustained reduction in population movement despite the relaxation of restrictions.

**Fig 1.**
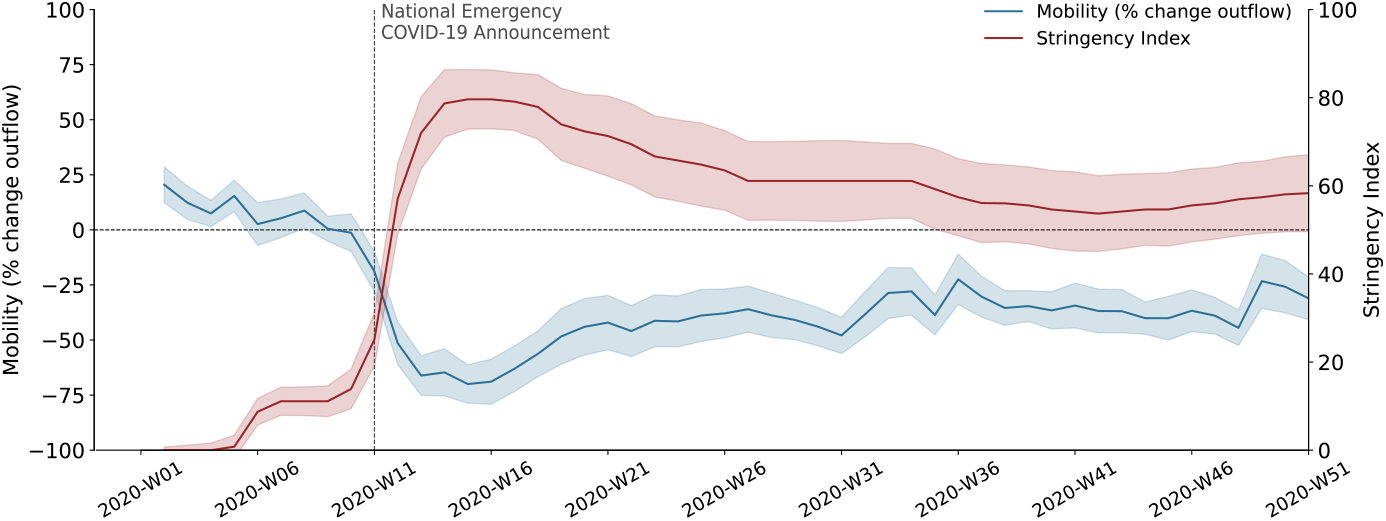
Mobility and policy stringency in the United States during 2020. Population mobility declined as policy stringency increased across U.S. states, with the strongest divergence following the national emergency declaration in week 11 of 2020. Solid lines show state-level averages, and shaded areas indicate one standard deviation around the mean.

The observed persistent reduction in mobility motivates the exploration of its relationship with self-reported mental health indicators. Unless otherwise specified, we restrict our analysis to April–September, to capture the first wave of infection. We explore the univariate rank correlation between the relative change in mobility and self-reported anxiety and depression levels across U.S. states. Using the Spearman correlation coefficient, we evaluate the strength and direction of these associations within each state. The resulting spatial patterns are displayed in Figure 2, which maps the state-level correlations between changes in mobility and mental health outcomes. Across the U.S., 38 out of 51 (nearly 75%) states show statistically significant correlations considering anxiety as mental health condition, and 31 out of 51 (nearly 61%) states considering depression. All significant states show negative coefficients considering anxiety, indicating higher anxiety associated with reduced mobility. A similar pattern has been observed considering depression, with the only exception of 1 out of 31 significant states showing a positive coefficient. The non-significant states are shown in white. It is important to note that these results are based on univariate regressions; therefore, the estimated correlations may reflect spurious associations arising from omitted variables.

**Fig 2.**
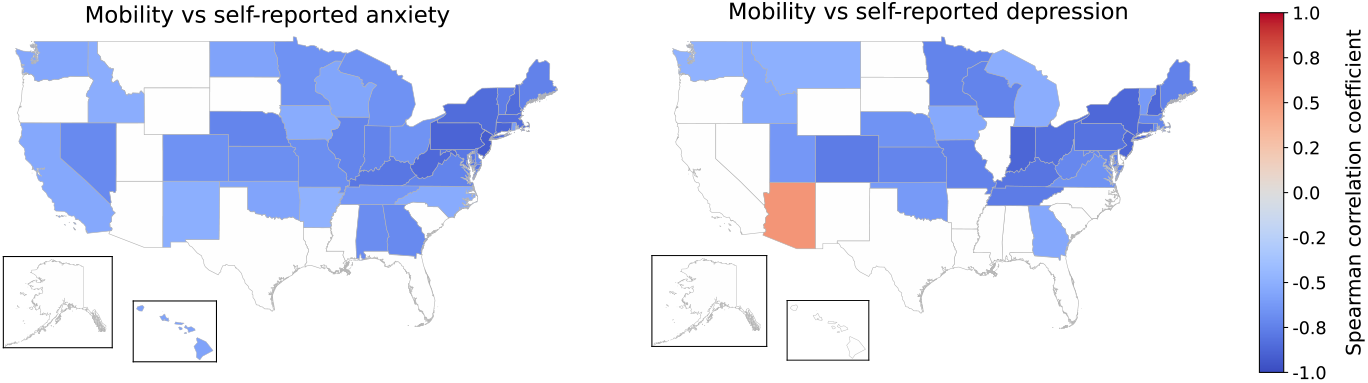
Spatial distribution of correlations between mobility and self-reported mental health. State-level Spearman correlation coefficients between relative mobility changes and self-reported anxiety (left) and depression (right). States with non-significant correlation coefficients (*p >* 0.05) are shown in white. The analysis is based on univariate regressions and does not control for potential confounders.

Guided by the state-level correlations, we further explore the multivariate association between mobility and self-reported mental health indicators through fixed effects panel regression models. Table 1 reports the results, with both models controlling for the COVID-19 policy stringency index and the mortality rate (log-transformed), while accounting for unobserved time-invariant state heterogeneity via entity fixed effects.

**Table 1.** Fixed effects regression of mobility on mental health indicators. Panel regression estimates with mobility change (relative to pre-pandemic baseline) as the dependent variable across 48 US states from April to September 2020. Model (1) examines the association with self-reported anxiety; Model (2) examines the association with self-reported depression. Both models control for policy stringency and COVID-19 mortality rates. All variables are standardized. State fixed effects are included. Standard errors are reported in parentheses.

|  | Mobility |  |
| --- | --- | --- |
|  | (1) Anxiety | (2) Depression |
| Stringency Index | −1.588**<br>(0.722) | −0.646<br>(0.931) |
| Mortality | 8.242***<br>(1.057) | 9.104***<br>(0.822) |
| Self-reported Anxiety | −7.195***<br>(1.034) |  |
| Self-reported Depression |  | −4.801***<br>(0.800) |
| Constant | −47.817***<br>(0.026) | −47.760***<br>(0.029) |
| Observations | 816 | 816 |
| States | 48 | 48 |
| R<sup>2</sup> (within) | 0.664 | 0.630 |

Notes: \* $p < 0.1$ ; \*\* $p < 0.05$ ; \*\*\* $p < 0.01$

In both configurations, the stringency index shows a significant negative association with mobility, indicating that stricter containment measures were consistently linked to reduced movement across states. The mortality rate exhibits a strong positive and significant coefficient, suggesting that states that reported higher COVID-19 mortality tended to experience higher mobility.

When including self-reported anxiety, we find a significant negative coefficient (*β*= *−*7.195, *p <* 0.01), implying that higher anxiety levels are associated with lower mobility outflow within states. Similarly, the model including self-reported depression yields a significant negative coefficient (*β*=*−*4.801, *p <* 0.01), suggesting that elevated self-reported depressive symptoms are also linked to reduced mobility.

Overall, the models explain a substantial proportion of within-state variation in mobility, supporting the robustness of the associations. Collectively, these findings suggest that both psychological distress and stricter policy interventions are associated with the sustained reductions in mobility observed during the COVID-19 pandemic’s first wave.

## Discussion

In this study, we conducted a statistical analysis to explain the variation of population-level mobility within U.S. states and territories in the aftermath of the COVID-19 national emergency announcement. Crossing state-level mobility data with longitudinal large-scale mental distress indicators collected with online surveys, we analysed associations between mobility recovery trends, self-reported feelings of anxiety and depression, stringency of interventions, and reported mortality.

Our findings suggest that higher mental distress had a significant association with lower mobility beyond the effect of stringency of restrictions or deaths. Specifically, our regression model suggests that, within states, population-level mobility was lower in weeks with higher mental distress levels despite equal stringency of interventions and number of deaths. The highest mental distress period was registered after the lockdown declaration (see Fig. *S*2, Supplementary Information). Similarly, the first relaxation of measures might have gradually reduced population mental distress levels, in the hope of a return to normality. Hence, in a first period of constant but milder restrictions compared to lockdown, lower mental distress may have induced a higher mobility recovery, not explained by reduced stringency, which is consistent with the loss of adherence to restrictions over time documented in previous studies [29]. Previous research [16] has pointed to risk adaptation as a dominant mechanism driving this loss of adherence to pandemic restrictions as a consequence of the psychological burden of prolonged isolation. Our approach does not prove nor discard that risk adaptation may have shaped loss of adherence for a fraction of the population, and it cannot be interpreted as a rejection of the risk adaptation hypothesis.

We found a significant positive association between reported COVID-19 mortality and mobility, such that states experiencing higher mortality tended to exhibit relatively higher mobility. This relationship cannot be read as a causal directionality, but rather as the result of two possible mechanisms. The observation that states with higher COVID-19 mortality did not exhibit proportionally larger reductions in mobility can suggest that the capacity to reduce mobility was limited even in high-risk contexts.

This pattern may reflect economic, social, or structural constraints, such as occupational requirements or limited opportunities for remote work, that restricted behavioral adaptation despite elevated mortality, as observed in other countries [37, 38]. While the interpretation based on higher mortality induces behavioral changes leading to reduced mobility cannot be ruled out, disentangling this mechanism is challenging without explicitly modeling temporal lags. This limitation is discussed further below. The positive association between mortality and mobility may reflect bidirectional mechanisms and delayed effects not fully captured by the current model. On the one hand, as the relaxation of measures unfolded, periods characterized by higher mobility levels were associated with increased COVID-19 deaths, consistent with a transmission-mediated pathway in which increased mobility leads to higher contact rates, greater case growth, and a delayed increase in mortality [39–41]. This is consistent with previous evidence showing that reductions in effective mobility were associated with lower case growth following the implementation of non-pharmaceutical interventions [42]. Given the persistence of infections over time, the impact of elevated mobility on mortality may extend beyond short temporal windows. On the other hand, behavioral responses to mortality may also operate over extended time scales. Previous studies have shown that directionality and significance of lagged relationships between mobility and epidemiological outcomes may vary across locations and epidemic waves, further highlighting the difficulty of identifying causal effects in this setting [43]. Given these complexities and in the absence of a causal inference framework, we focus on contemporaneous associations to explore this relationship at the population level.

This study comes with the following limitations. First, we rely on self-reported depression and anxiety, which do not correspond to clinical assessment or formal diagnosis and may be influenced by reporting biases. Second, we focus on a binary outcome, without considering intensity, duration, and the degrees or the nuances of anxiety and depression experienced by individuals. Third, our analysis does not capture dynamic population-level factors such as evolving social norms, stigma, or fear of legal repercussions associated with non-compliance to mobility restrictions, which may influence both behavior and mental health. Fourth, although we identify statistically significant associations between changes in mental distress and mobility recovery, our cross-sectional design does not allow causal inference and is not intended to disentangle bidirectional mechanisms. Additionally, our study does not account for lagged effects; it follows the framework of previous studies examining contemporaneous relationships between mobility and mental health [44], as well as between stringency, mental health, and mortality at the population level [45].

Overall, our findings highlight the importance of accounting for population mental health when interpreting behavioral responses to public health interventions. Mobility patterns during the pandemic were not only driven by policy stringency or epidemiological severity, but could also be shaped by collective psychological states. This suggests that policies aimed at controlling disease spread may benefit from being accompanied by interventions addressing mental distress, such as transparent risk communication, social support measures, and accessible mental health services. A more comprehensive understanding of the interplay between psychological well-being and collective behavior is needed to design effective and socially sustainable responses to future public health emergencies.

## Data Availability

All data produced are described and available in the following references. The mobility data are described in: Kang, Y., Gao, S., Liang, Y. Li, M., Rao, J. and Kruse, J. Multiscale dynamic human mobility flow dataset in the U.S. during the COVID-19 epidemic. Scientific Data 7, 390 (2020). https://www.nature.com/articles/s41597-020-00734-5. The Github repository containing the data is available: https://github.com/GeoDS/COVID19USFlows-WeeklyFlows
The mental health survey data are presented in: Salomon JA, Reinhart A, Bilinski A, Chua EJ, La Motte-Kerr W, Ronn MM, et al. The US COVID-19 Trends and Impact Survey: Continuous real-time measurement of COVID-19 symptoms, risks, protective behaviors, testing, and vaccination. Proceedings of the National Academy of Sciences. 2021;118(51):e2111454118. Access can be requested online (https://cmu-delphi.github.io/delphi-epidata/symptom-survey/data-access.html)

https://cmu-delphi.github.io/delphi-epidata/symptom-survey/data-access.html

## Acknowledgements

SF, SK, PC, SG, SMP, FM, ADG, DL, JDI, KK, MT and MM acknowledge support from the Lagrange Project of the ISI Foundation, funded by Fondazione CRT.

