## Supplementary Information for "Characterising associations between mental distress, mobility, and COVID-19 restrictions: a U.S. study"

Stefania Fiandrino[1,2], Saumitra Kulkarni[2], Paolo Cornale[2], Sara Ghivarello[2], Piero Birello[3], Simone Maria Parazzoli[2], Federico Moss[2], Alessandro De Gaetano[2], Daniele Liberatore[2], Jacopo D'Ignazi[2], Kyriaki Kalimeri[2], Michele Tizzani[2,4], Mattia Mazzoli[2]

[1] Department of Computer, Control, and Management Engineering Antonio Ruberti, Sapienza University of Rome, Rome, Italy

[2] ISI Foundation, Turin, Italy

[3] Politecnico di Torino, Turin, Italy

[4] DTU, Copenhagen, Denmark

### List of Figures

S1 **Pairwise relationships among mobility, self-reported mental health indicators, policy stringency, and mortality.** Diagonal panels show marginal distributions for each variable. Off-diagonal panels display bivariate density estimates in grayscale, where darker shades indicate higher observation density. . . . . . . . . . . . . . . . . . . . . . . . . . . . . . . . . 3

S2 **Weekly trends in mobility and self-reported mental health across U.S. states in 2020.** The blue line shows the weekly percentage change in outflow mobility relative to baseline. Green and orange lines report the reported symptoms of depression and anxiety, respectively. Shaded bands indicate the confidence interval across states. The vertical dashed line marks the national COVID-19 emergency declaration. . . . . . . . . . . . . . . . . . . . . . 4

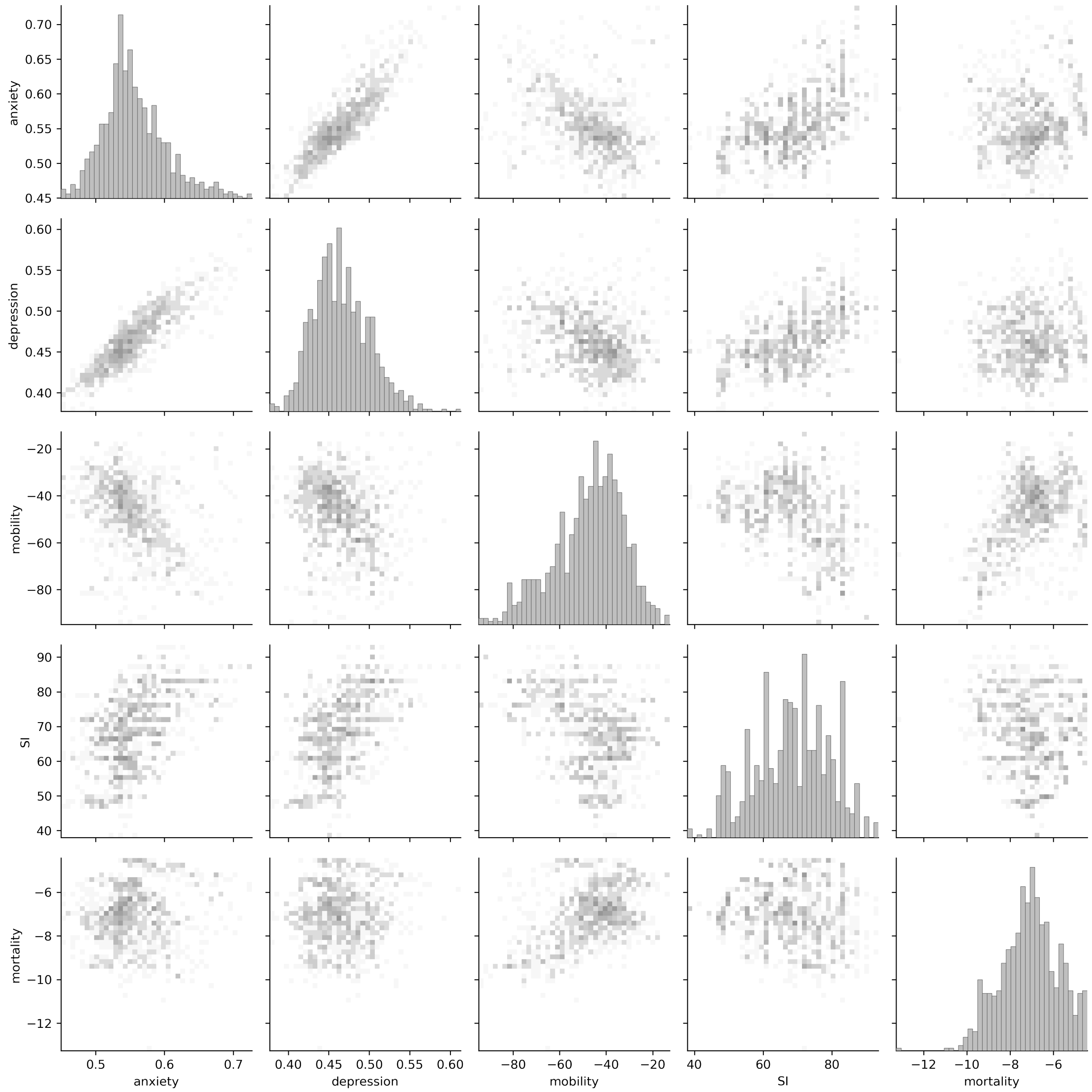


Figure S1: **Pairwise relationships among mobility, self-reported mental health indicators, policy stringency, and mortality.** Diagonal panels show marginal distributions for each variable. Off-diagonal panels display bivariate density estimates in grayscale, where darker shades indicate higher observation density.

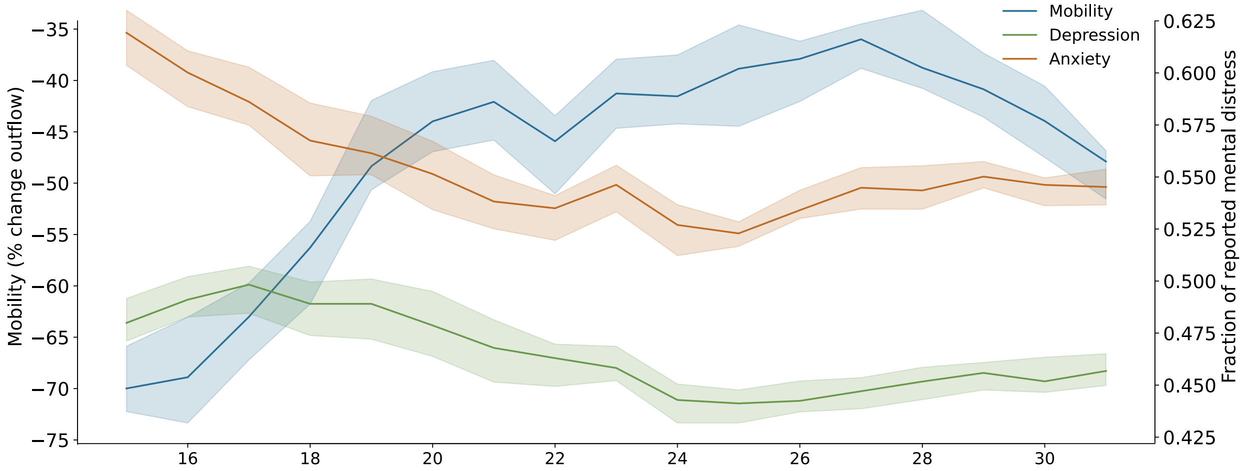


Figure S2: **Weekly trends in mobility and self-reported mental health across U.S. states in 2020.** The blue line shows the weekly percentage change in outflow mobility relative to baseline. Green and orange lines report the reported symptoms of depression and anxiety, respectively. Shaded bands indicate the confidence interval across states. The vertical dashed line marks the national COVID-19 emergency declaration.
